# Effect of Hydroxychloroquine in Hospitalized Patients with COVID-19: Preliminary results from a multi-centre, randomized, controlled trial

**DOI:** 10.1101/2020.07.15.20151852

**Authors:** Peter Horby, Marion Mafham, Louise Linsell, Jennifer L Bell, Natalie Staplin, Jonathan R Emberson, Martin Wiselka, Andrew Ustianowski, Einas Elmahi, Benjamin Prudon, Anthony Whitehouse, Timothy Felton, John Williams, Jakki Faccenda, Jonathan Underwood, J Kenneth Baillie, Lucy Chappell, Saul N Faust, Thomas Jaki, Katie Jeffery, Wei Shen Lim, Alan Montgomery, Kathryn Rowan, Joel Tarning, James A Watson, Nicholas J White, Edmund Juszczak, Richard Haynes, Martin J Landray

## Abstract

**Background:** Hydroxychloroquine and chloroquine have been proposed as treatments for coronavirus disease 2019 (COVID-19) on the basis of in vitro activity, uncontrolled data, and small randomized studies.

**Methods:** The Randomised Evaluation of COVID-19 therapy (RECOVERY) trial is a randomized, controlled, open-label, platform trial comparing a range of possible treatments with usual care in patients hospitalized with COVID-19. We report the preliminary results for the comparison of hydroxychloroquine vs. usual care alone. The primary outcome was 28-day mortality.

**Results:** 1561 patients randomly allocated to receive hydroxychloroquine were compared with 3155 patients concurrently allocated to usual care. Overall, 418 (26.8%) patients allocated hydroxychloroquine and 788 (25.0%) patients allocated usual care died within 28 days (rate ratio 1.09; 95% confidence interval [CI] 0.96 to 1.23; P=0.18). Consistent results were seen in all pre-specified subgroups of patients. Patients allocated to hydroxychloroquine were less likely to be discharged from hospital alive within 28 days (60.3% vs. 62.8%; rate ratio 0.92; 95% CI 0.85-0.99) and those not on invasive mechanical ventilation at baseline were more likely to reach the composite endpoint of invasive mechanical ventilation or death (29.8% vs. 26.5%; risk ratio 1.12; 95% CI 1.01-1.25). There was no excess of new major cardiac arrhythmia.

**Conclusions:** In patients hospitalized with COVID-19, hydroxychloroquine was not associated with reductions in 28-day mortality but was associated with an increased length of hospital stay and increased risk of progressing to invasive mechanical ventilation or death.

**Funding:** Medical Research Council and NIHR (Grant ref: MC_PC_19056).

**Trial registrations:** The trial is registered with ISRCTN (50189673) and clinicaltrials.gov (NCT04381936).

## INTRODUCTION

Severe acute respiratory syndrome coronavirus 2 (SARS-CoV-2), the cause of coronavirus disease 2019 (COVID-19), emerged in China in late 2019 from a zoonotic source.^1^ The majority of COVID-19 infections are either asymptomatic or result in only mild disease. However, a substantial proportion of infected individuals develop a respiratory illness requiring hospital care,^2^ which can progress to critical illness with hypoxemic respiratory failure requiring prolonged ventilatory support.^3-6^ Amongst COVID-19 patients admitted to UK hospitals, the case fatality rate is around 26%, and is over 37% in patients requiring invasive mechanical ventilation.^7^

Hydroxychloroquine and chloroquine, 4-aminoquinoline drugs developed over 70 years ago and used to treat malaria and rheumatological conditions, have been proposed as treatments for COVID-19. Chloroquine has in vitro activity against a variety of viruses, including SARS-CoV-2 and the related SARS-CoV-1.^8-13^ The exact mechanism of antiviral action is uncertain but these drugs increase the pH of endosomes that the virus uses for cell entry and also interfere with the glycosylation of the cellular receptor of SARS-CoV, angiotensin-converting enzyme 2 (ACE2), and associated gangliosides.^10,14^ The 4-aminoquinoline concentrations required to inhibit SARS-CoV-2 replication in vitro are relatively high by comparison with the free plasma concentrations observed in the prevention and treatment of malaria.^15^ These drugs are generally well tolerated, inexpensive and widely available. Following oral administration they are rapidly absorbed, even in severely ill patients. If active, therapeutic hydroxychloroquine concentrations could be expected in the human lung shortly after an initial loading dose.

Small pre-clinical studies have reported that hydroxychloroquine prophylaxis or treatment had no beneficial effect of clinical disease or viral replication.^16^ Clinical benefit and antiviral effect from the administration of these drugs alone or in combination with azithromycin to patients with COVID-19 infections has been reported in some observational studies ^17-21^ but not in others.^22-24^A few small controlled trials of hydroxychloroquine and chloroquine for the treatment of COVID-19 infection have been inconclusive.^25-28^ Here we report preliminary results of the effects of a randomized controlled trial of hydroxychloroquine in patients hospitalized with COVID-19.

## METHODS

### Trial design and participants

The RECOVERY trial is an investigator-initiated, individually randomized, controlled, open-label, platform trial to evaluate the effects of potential treatments in patients hospitalized with COVID-19. The trial is conducted at 176 hospitals in the United Kingdom (see Supplementary Appendix), supported by the National Institute for Health Research Clinical Research Network. The trial is coordinated by the Nuffield Department of Population Health at University of Oxford, the trial sponsor. Although the hydroxychloroquine, dexamethasone, and lopinavir-ritonavir arms have now been stopped, the trial continues to study the effects of azithromycin, tocilizumab, and convalescent plasma (and other treatments may be studied in the future).

Hospitalized patients were eligible for the study if they had clinically suspected or laboratory confirmed SARS-CoV-2 infection and no medical history that might, in the opinion of the attending clinician, put the patient at significant risk if they were to participate in the trial. Initially, recruitment was limited to patients aged at least 18 years but from 9 May 2020, the age limit was removed. Patients with known prolonged electrocardiograph QTc interval were ineligible for the hydroxychloroquine arm. Co-administration with medications that prolong the QT interval was not an absolute contraindication but attending clinicians were advised to check the QT interval by performing an electrocardiogram.

Written informed consent was obtained from all patients or from a legal representative if they were too unwell or unable to provide consent. The trial was conducted in accordance with the principles of the International Conference on Harmonization–Good Clinical Practice guidelines and approved by the UK Medicines and Healthcare Products Regulatory Agency (MHRA) and the Cambridge East Research Ethics Committee (ref: 20/EE/0101). The protocol and statistical analysis plan are available in the Supplementary Appendix and on the study website www.recoverytrial.net.

### Randomization

Baseline data collected using a web-based case report form included demographics, level of respiratory support, major comorbidities, the suitability of the study treatment for a particular patient, and treatment availability at the study site. Eligible and consenting patients were assigned in a ratio of 2:1 to either usual standard of care or usual standard of care plus hydroxychloroquine or one of the other available treatment arms (see Supplementary Appendix) using web-based simple (unstratified) randomization with allocation concealment. Patients allocated to hydroxychloroquine sulfate (200mg tablet containing 155mg base equivalent) received a loading dose of 4 tablets (800 mg) at zero and 6 hours, followed by 2 tablets (400 mg) starting at 12 hours after the initial dose and then every 12 hours for the next 9 days or until discharge (whichever occurred earlier) (see Supplementary Appendix).^15^ Allocated treatment was prescribed by the attending clinician. Participants and local study staff were not blinded to the allocated treatment.

### Procedures

A single online follow-up form was to be completed when participants were discharged, had died or at 28 days after randomization (whichever occurred earlier). Information was recorded on adherence to allocated study treatment, receipt of other study treatments, duration of admission, receipt of respiratory support (with duration and type), receipt of renal dialysis or hemofiltration, and vital status (including cause of death). From 12 May 2020, extra information was recorded on the occurrence of new major cardiac arrhythmia. In addition, routine health care and registry data were obtained including information on vital status (with date and cause of death); discharge from hospital; respiratory and renal support therapy.

### Outcome measures

Outcomes were assessed at 28 days after randomization, with further analyses specified at 6 months. The primary outcome was all-cause mortality. Secondary outcomes were time to discharge from hospital and, among patients not on invasive mechanical ventilation at randomization, invasive mechanical ventilation (including extra-corporal membrane oxygenation) or death. Subsidiary clinical outcomes included cause-specific mortality, use of hemodialysis or hemofiltration, major cardiac arrhythmia (recorded in a subset), and receipt and duration of ventilation.

### Statistical Analysis

For the primary outcome of 28-day mortality, the log-rank ‘observed minus expected’ statistic and its variance were used to both test the null hypothesis of equal survival curves and to calculate the one-step estimate of the average mortality rate ratio, comparing all patients allocated hydroxychloroquine with all patients allocated usual care. The few patients (2.1%) who had not been followed for 28 days by the time of the data cut (22 June 2020) were either censored on 22 June 2020 or, if they had already been discharged alive, were right-censored for mortality at day 29 (that is, in the absence of any information to the contrary they were assumed to have survived 28 days). Kaplan-Meier survival curves were constructed to display cumulative mortality over the 28-day period. The same methods were used to analyze time to hospital discharge, with patients who died in hospital right-censored on day 29. Median time to discharge was derived from the Kaplan-Meier estimates. For the pre-specified composite secondary outcome of invasive mechanical ventilation or death within 28 days (among those not receiving invasive mechanical ventilation at randomization), the precise date of starting invasive mechanical ventilation was not available and so the risk ratio was estimated instead. Estimates of absolute risk differences between patients allocated hydroxychloroquine and patients allocated usual care were also calculated.

Pre-specified analyses of the primary outcome were performed in five subgroups defined by characteristics at randomization: age, sex, level of respiratory support, days since symptom onset, and predicted 28-day mortality risk (See Supplementary Appendix). One further pre-specified subgroup analysis (ethnicity) will be conducted once data collection is completed. Observed effects within subgroup categories were compared using a chi-square test for trend (which is equivalent to a test for heterogeneity for subgroups that have only two levels).

Estimates of rate and risk ratios (both hereon denoted RR) are shown with 95% confidence intervals. All p-values are 2-sided and are shown without adjustment for multiple testing. All analyses were done according to the intention-to-treat principle. The full database is held by the study team which collected the data from study sites and performed the analyses at the Nuffield Department of Population Health, University of Oxford.

### Sample size and decision to stop enrolment

As stated in the protocol, appropriate sample sizes could not be estimated when the trial was being planned at the start of the COVID-19 pandemic. As the trial progressed, the Trial Steering Committee, blinded to the results of the study treatment comparisons, formed the view that if 28-day mortality was 20% then a comparison of at least 2000 patients allocated to active drug and 4000 to usual care alone would yield at least 90% power at two-sided P=0.01 to detect a proportional reduction of one-fifth (a clinically relevant absolute difference of 4 percentage points between the two arms).

The independent Data Monitoring Committee reviewed unblinded analyses of the study data and any other information considered relevant at intervals of around 2 weeks. The committee was charged with determining if, in their view, the randomized comparisons in the study provided evidence on mortality that is strong enough (with a range of uncertainty around the results that is narrow enough) to affect national and global treatment strategies. In such a circumstance, the Committee would inform the Trial Steering Committee who would make the results available to the public and amend the trial arms accordingly. Unless that happened, the Trial Steering Committee, investigators, and all others involved in the trial would remain blind to the interim results until 28 days after the last patient had been randomized to a particular intervention arm.

On 4 June, in response to a request from the MHRA, the independent Data Monitoring Committee conducted a review of the data and recommended the chief investigators review the unblinded data on the hydroxychloroquine arm of the trial. The Chief Investigators and Trial Steering Committee concluded that the data showed no beneficial effect of hydroxychloroquine in patients hospitalized with COVID-19. Therefore enrolment of participants to the hydroxychloroquine arm was closed on 5 June and the preliminary result for the primary outcome was made public. Investigators were advised that any patients currently taking hydroxychloroquine as part of the study should discontinue the treatment.

## RESULTS

### Patients

Of the 11,197 patients randomized while the hydroxychloroquine arm was open (25 March to 5 June 2020), 7513 (67%) were eligible to be randomized to hydroxychloroquine (that is hydroxychloroquine was available in the hospital at the time and the attending clinician was of the opinion that the patient had no known indication for or contraindication to hydroxychloroquine) (Figure 1 and Table S1). Of these, 1561 were randomized to hydroxychloroquine and 3155 were randomized to usual care with the remainder being randomized to one of the other treatment arms. Mean age of study participants in this comparison was 65.3 (SD 15.3) years (Table 1) and 38% patients were female. No children were enrolled in the hydroxychloroquine comparison. A history of diabetes was present in 27% of patients, heart disease in 26%, and chronic lung disease in 22%, with 57% having at least one major comorbidity recorded. In this analysis, 90% of patients had laboratory confirmed SARS-CoV-2 infection, with the result currently awaited for 1%. At randomization, 17% were receiving invasive mechanical ventilation or extracorporeal membrane oxygenation, 60% were receiving oxygen only (with or without non-invasive ventilation), and 24% were receiving neither.

**Table 1:**
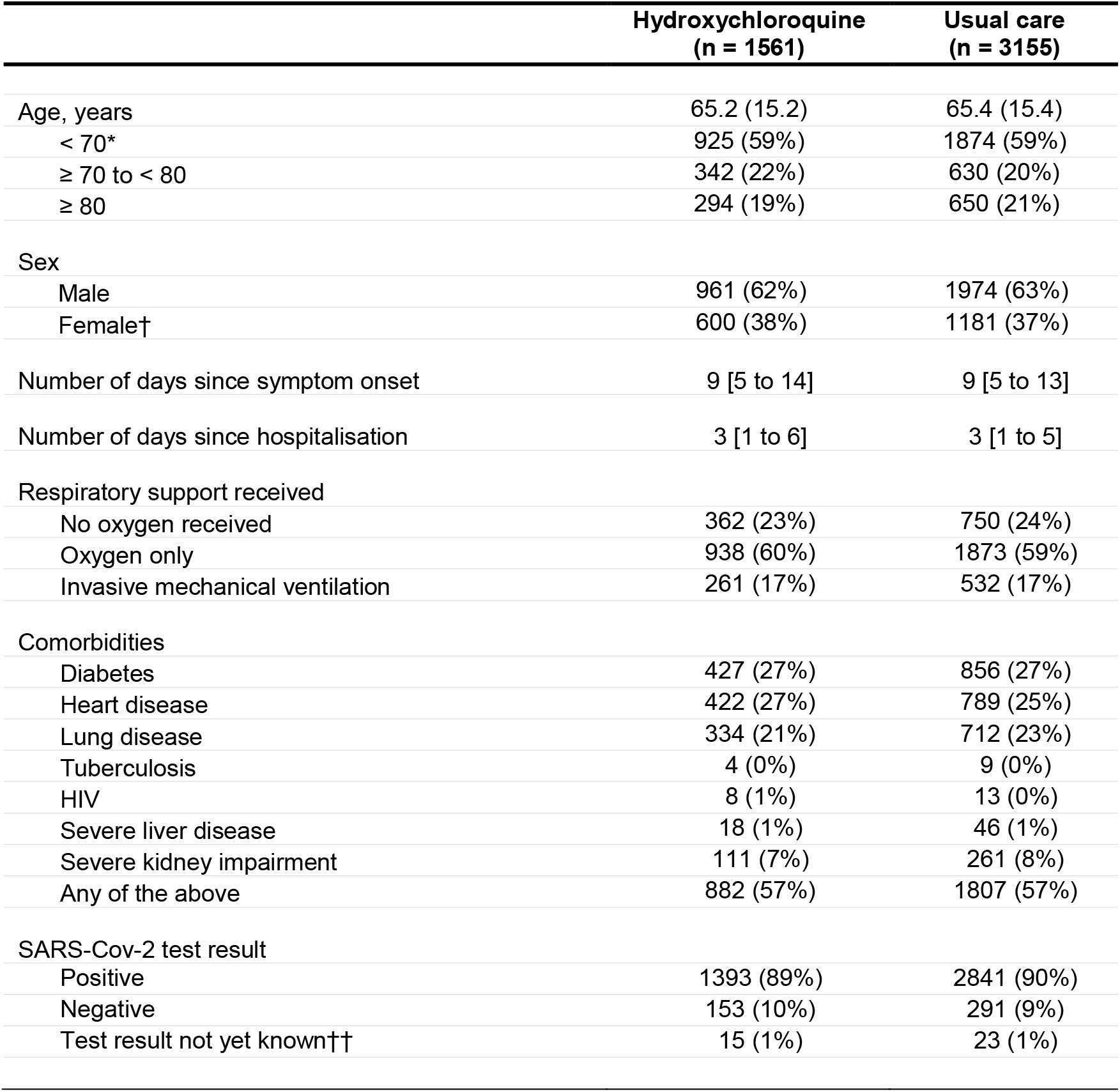
Baseline characteristics by randomized allocation. Results are count (%), mean ± standard deviation, or median (inter-quartile range).* No children (aged <18 years) were enrolled. †Includes 6 pregnant women. †† SARS-Cov-2 test results are captured on the follow-up form, so are currently unknown for some. All tests for difference in baseline characteristics between treatment arms give p>0.05. The ‘oxygen only’ group includes non-invasive ventilation. Severe liver disease defined as requiring ongoing specialist care. Severe kidney impairment defined as estimated glomerular filtration rate <30 mL/min/1.73m^2^. 9 (0.6%) patients allocated to hydroxychloroquine and 9 (0.3%) patients allocated to usual care alone had missing data for days since symptom onset.

**Figure 1:**
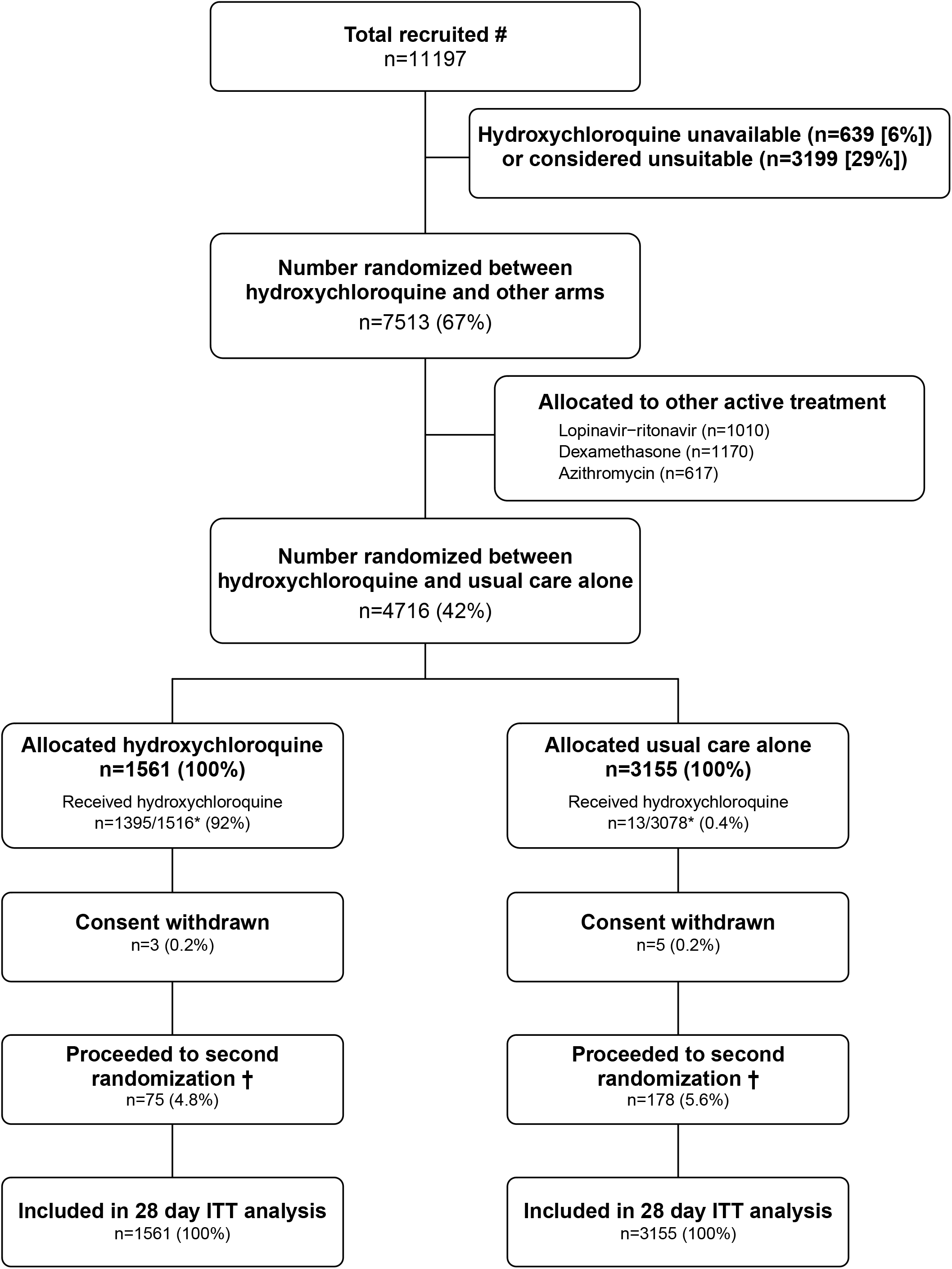
Trial profile - Flow of participants through the RECOVERY trial. ITT=intention to treat. * Number recruited overall during period that adult participants could be recruited into hydroxychloroquine comparison. # 1516/1561 (97.1%) and 3078/3155 (97.6%) patients have a completed follow-up form at time of analysis. † includes 37/1561 (2.4%) patients in the hydroxychloroquine arm and 89/3155 (2.8%) patients in the usual care arm allocated to tocilizumab in accordance with protocol version 4.0 or later. 6 patients were additionally randomized to convalescent plasma vs control (1 [0.1%] patient allocated to hydroxychloroquine arm vs 5 [0.2%] patients allocated to usual care) in accordance with protocol version 6.0. Among the 167 sites that randomized at least 1 patient to the hydroxychloroquine comparison, the median number randomized was 20 patients (inter-quartile range 11 to 41).

Follow-up information was complete for 4619 (98%) of the randomized patients. Among those with a completed follow-up form, 1395 (92%) patients allocated to hydroxychloroquine received at least 1 dose (Table S2) and the median number of days of treatment was 6 days (IQR 3 to 10 days). 13 (0.4%) of the usual care arm received hydroxychloroquine. Use of azithromycin or other macrolide drug during the follow-up period was similar in both arms (17% vs. 19%) as was use of dexamethasone (8% vs. 9%).

### Primary outcome

There was no significant difference in the proportion of patients who met the primary outcome of 28-day mortality between the two randomized arms (418 [26.8%] patients in the hydroxychloroquine arm vs. 788 [25.0%] patients in the usual care arm; rate ratio, 1.09; 95% confidence interval [CI], 0.96 to 1.23; P=0.18) (Figure 2). Similar results were seen across all five pre-specified subgroups (Figure 3). In post hoc exploratory analyses restricted to the 4234 (90%) patients with a positive SARS-CoV-2 test result, the result was similar (rate ratio, 1.09, 95% CI 0.96 to 1.24).

**Figure 2:**
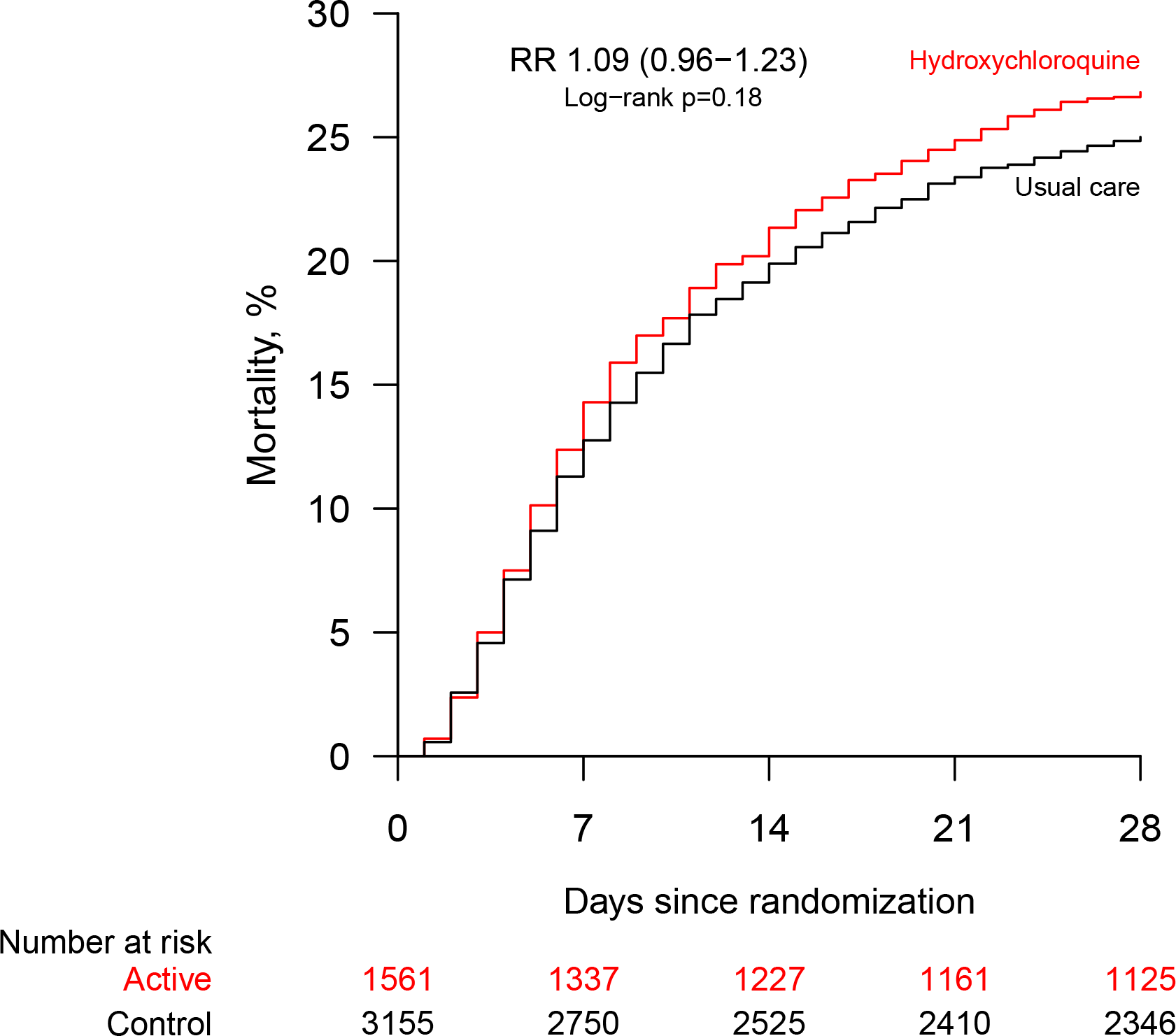
28−day mortality. RR=rate ratio. CI=confidence interval. The RR is derived from the log-rank observed minus expected statistic (O – E) and its variance (V) as the one-step estimate, through the formula exp([O – E] ÷ V), and its 95% CI is given by exp([O – E] ÷ V ± 1.96 ÷ √V). The number of patients randomized and the number remaining at risk of death at the end of days 7, 14, 21 and 28 are shown beneath the plot.

**Figure 3:**
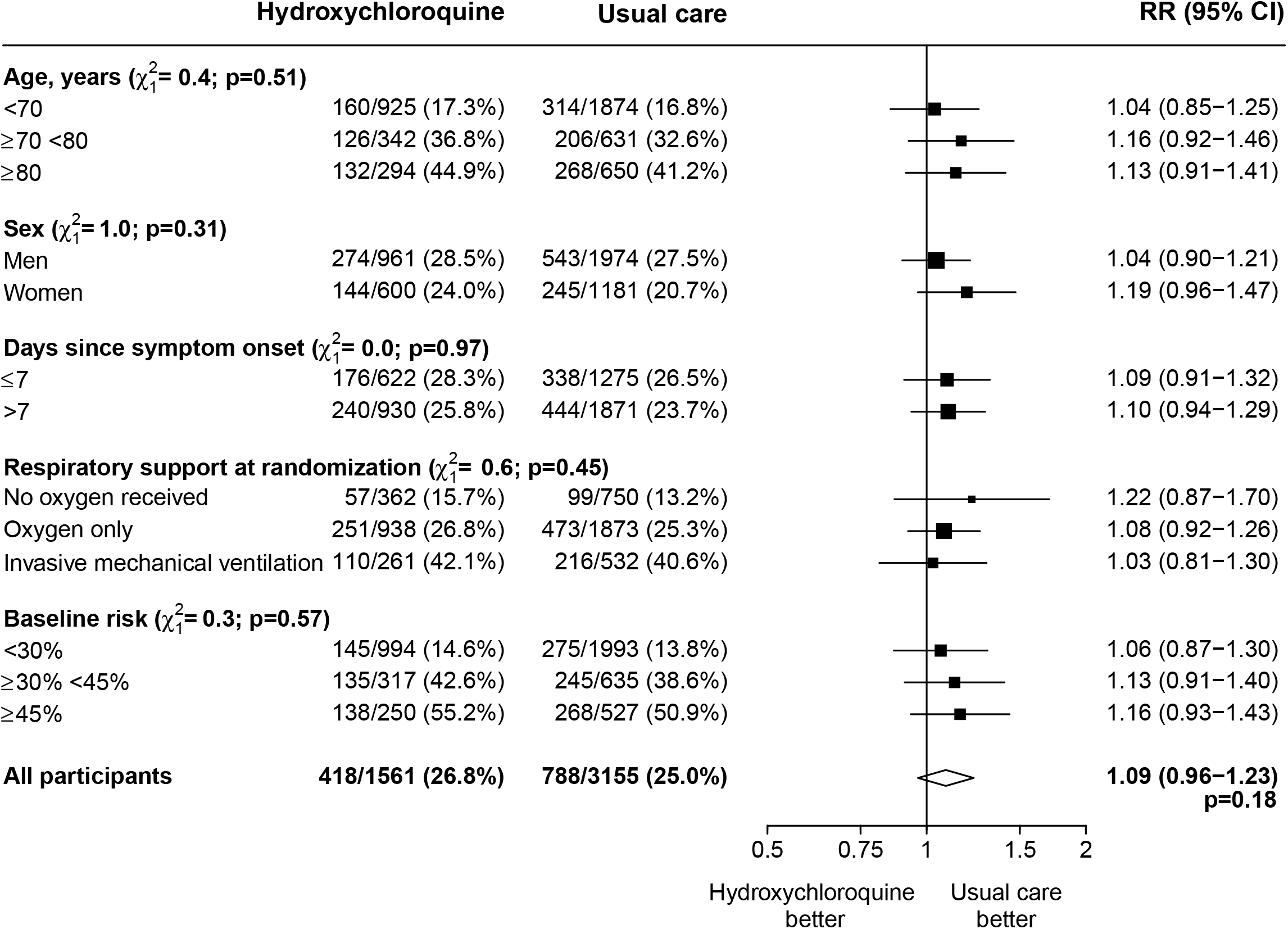
Effect of allocation to hydroxychloroquine on 28-day mortality by pre-specified characteristics at randomization. RR=rate ratio. CI=confidence interval. Subgroup−specific RR estimates are represented by squares (with areas of the squares proportional to the amount of statistical information) and the lines through them correspond to the 95% confidence intervals. The ‘oxygen only’ group includes patients receiving non-invasive ventilation. The method used for calculating baseline-predicted risk is described in the Supplementary Appendix. One further pre-specified subgroup analysis (ethnicity) will be conducted once data collection is completed.

### Secondary outcomes

Allocation to hydroxychloroquine was associated with a longer time until discharge alive from hospital than usual care (median 16 days vs. 13 days) and a lower probability of discharge alive within 28 days (rate ratio 0.92, 95% CI 0.85 to 0.99) (Table 2). Among those not on invasive mechanical ventilation at baseline, the number of patients progressing to the pre-specified composite secondary outcome of invasive mechanical ventilation or death was higher among those allocated to hydroxychloroquine (risk ratio 1.12, 95% CI 1.01 to 1.25).

**Table 2:** Effect of allocation to hydroxychloroquine on main study outcomes. RR=rate ratio for the outcomes of 28-day mortality and hospital discharge, and risk ratio for the outcome of receipt of invasive mechanical ventilation or death. CI=confidence interval. * Analyses exclude those on invasive mechanical ventilation at randomization. For the pre-specified composite secondary endpoint of receipt of invasive mechanical ventilation or death the absolute risk difference was 3.3 percentage points (95% CI 0.3 to 6.3).

|  | Hydroxychloroquine<br>(n = 1561) | Usual care<br>(n = 3155) | RR<br>(95% CI) |
| --- | --- | --- | --- |
| **Primary outcome:** |  |  |  |
| 28-day all-cause mortality | 418 (26.8%) | 788 (25.0%) | 1.09 (0.96 to 1.23) |
| **Secondary outcomes:** |  |  |  |
| Discharged from hospital within 28 days | 941 (60.3%) | 1982 (62.8%) | 0.92 (0.85 to 0.99) |
| Receipt of mechanical ventilation or death* | 388/1300 (29.8%) | 696/2623 (26.5%) | 1.12 (1.01 to 1.25) |
| Death | 308/1300 (23.7%) | 572/2623 (21.8%) | 1.09 (0.96 to 1.23) |
| Invasive mechanical ventilation | 118/1300 (9.1%) | 215/2623 (8.2%) | 1.11 (0.89 to 1.37) |

### Subsidiary outcomes

Information on the occurrence of new major cardiac arrhythmia was collected for 698 (44.7%) patients in the hydroxychloroquine arm and 1357 (43.0%) in the usual care arm since these fields were added to the follow-up form on 12 May 2020. Among these patients, there were no significant differences in the frequency of supraventricular tachycardia (6.9% vs. 5.9%), ventricular tachycardia or fibrillation (0.9% vs. 0.7%) or atrioventricular block requiring intervention (0.1% vs. 0.1%) (Table S3). Analyses of cause-specific mortality, receipt of renal dialysis or hemofiltration, and duration of ventilation will be presented once all relevant information (including certified cause of death) is available. There was one report of a serious adverse reaction believed related to hydroxychloroquine; a case of torsades de pointes from which the patient recovered without the need for intervention.

## DISCUSSION

Although preliminary, these results indicate that hydroxychloroquine is not an effective treatment for patients hospitalized with COVID-19. The lower bound of the confidence limit for the primary outcome rules out any reasonable possibility of a meaningful mortality benefit. In addition, allocation to hydroxychloroquine was associated with an increase in the duration of hospitalization and an increased risk of requiring invasive mechanical ventilation or dying for those not on invasive mechanical ventilation at baseline. The results were consistent across subgroups of age, sex, time since illness onset, level of respiratory support, and baseline-predicted risk.

RECOVERY is a large, pragmatic, randomized, controlled platform trial designed to provide rapid and robust assessment of the impact of readily available potential treatments for COVID-19 on 28-day mortality. Around 15% of all patients hospitalized with COVID-19 in the UK over the study period were enrolled in the trial and the fatality rate in the usual care arm is consistent with the hospitalized case fatality rate in the UK and elsewhere.^7,29,30^ Only essential data were collected at hospital sites with additional information (including long-term mortality) ascertained through linkage with routine data sources. We did not collect information on physiological, electrocardiographic, laboratory or virologic parameters.

Hydroxychloroquine has been proposed as a treatment for COVID-19 based largely on its *in vitro* SARS-CoV-2 antiviral activity and on data from observational studies reporting effective reduction in viral loads. However, the 4-aminoquinoline drugs are relatively weak antivirals.^15^ Demonstration of therapeutic efficacy of hydroxychloroquine in severe COVID-19 would require rapid attainment of efficacious levels of free drug in the blood and respiratory epithelium.^31^ Thus, to provide the greatest chance of providing benefit in life threatening COVID-19, the dose regimen was designed to result in rapid attainment and maintenance of plasma concentrations that were as high as safely possible.^15^ These concentrations were predicted to be at the upper end of those observed during steady state treatment of rheumatoid arthritis with hydroxychloroquine.^32^ Our dosing schedule was based on hydroxychloroquine pharmacokinetic modelling referencing a SARS-CoV-2 half maximal effective concentration (EC_50_) of 0.72 μM scaled to whole blood concentrations and an assumption that cytosolic concentrations in the respiratory epithelium are in dynamic equilibrium with blood concentrations.^8,15,33^

The primary concern with short-term high dose 4-aminoquinoline regimens is cardiovascular toxicity. Hydroxychloroquine causes predictable prolongation of the electrocardiograph QT interval that is exacerbated by co-administration with azithromycin, as widely prescribed in COVID-19 treatment.^16-18^ Although torsade de pointes has been described, serious cardiovascular toxicity has been reported very rarely despite the high prevalence of cardiovascular disease in hospitalized patients, the common occurrence of myocarditis in COVID-19, and the extensive use of hydroxychloroquine and azithromycin together. The exception is a Brazilian study which was stopped early because of cardiotoxicity. However in that study, chloroquine 600 mg base was given twice daily for ten days, a substantially higher total dose than used in other trials, including RECOVERY.^34,35^ Pharmacokinetic modelling in combination with blood concentration and mortality data from a case series of 302 chloroquine overdose patients predicts that the base equivalent chloroquine regimen to the RECOVERY hydroxychloroquine regimen is safe.^35^ Hydroxychloroquine is considered to be safer than chloroquine.^15^ We did not observe excess mortality in the first 2 days of treatment with hydroxychloroquine, the time when early effects of dose-dependent toxicity might be expected. Furthermore, the preliminary data presented here did not show any excess in ventricular tachycardia (including torsade de pointes) or ventricular fibrillation in the hydroxychloroquine arm.

The findings indicate that hydroxychloroquine is not an effective treatment for hospitalized patients with COVID-19 but do not address its use as prophylaxis or in patients with less severe SARS-CoV-2 infection managed in the community. Treatment of COVID-19 with chloroquine or hydroxychloroquine has been recommended in many treatment guidelines, including in Brazil, China, France, Italy, Netherlands, South Korea, and the United States.^36^ In a retrospective cohort study in the United States, 59% of 1376 COVID-19 patients received hydroxychloroquine.^22,37^ Since our preliminary results were first made public on 5 June 2020, the U.S. Food and Drugs Administration has revoked the Emergency Use Authorization that allowed hydroxychloroquine and chloroquine to be used for hospitalized patients with COVID-19,^38^ and the World Health Organization (WHO) and the National Institutes for Health have ceased trials of its use in this setting on the grounds of lack of benefit. The WHO has recently released preliminary results from the SOLIDARITY trial on the effectiveness of hydroxychloroquine in hospitalized COVID-19 patients that are consistent with the results from the RECOVERY trial.^39^

### Authorship

This manuscript was initially drafted by the first and last author, developed by the Writing Committee, and approved by all members of the Trial Steering Committee. The funders had no role in the analysis of the data, preparation and approval of this manuscript, or the decision to submit it for publication. The first and last members of the Writing Committee vouch for the data and analyses, and for the fidelity of this report to the study protocol and data analysis plan.

### Writing Committee (on behalf of the RECOVERY Collaborative Group)

Peter Horby FRCP,^a,^* Marion Mafham MD,^b,*^ Louise Linsell DPhil,^b,*^ Jennifer L Bell MSc,^b^ Natalie Staplin PhD,^b,c^ Jonathan R Emberson PhD,^b,c^ Martin Wiselka PhD,^d^ Andrew Ustianowski PhD,^e^ Einas Elmahi MPhil,^f^ Benjamin Prudon FRCP,^g^ Anthony Whitehouse FRCA,^h^ Timothy Felton PhD,^i^ John Williams MRCP,^j^ Jakki Faccenda MD,^k^ Jonathan Underwood PhD,^l^ J Kenneth Baillie MD PhD,^m^ Lucy C Chappell PhD,^n^ Saul N Faust FRCPCH,° Thomas Jaki PhD,^p,q^ Katie Jeffery PhD,^r^ Wei Shen Lim FRCP,^s^ Alan Montgomery PhD,^t^ Kathryn Rowan PhD,^u^ Joel Tarning PhD,^v,w^ James A Watson DPhil,^v,w^ Nicholas J White FRS,^v,w^ Edmund Juszczak MSc,^b,†^ Richard Haynes DM,^b,c,†^ Martin J Landray PhD.^b,c,x,†^

^a^ Nuffield Department of Medicine, University of Oxford, Oxford, United Kingdom.

^b^ Nuffield Department of Population Health, University of Oxford, Oxford, United Kingdom

^c^ MRC Population Health Research Unit, University of Oxford, Oxford, United Kingdom

^d^ University Hospitals of Leicester NHS Trust and University of Leicester, Leicester, United Kingdom

^e^ Regional Infectious Diseases Unit, North Manchester General Hospital & University of Manchester, Manchester, United Kingdom

^f^ Research and Development Department, Northampton General Hospital, Northampton, United Kingdom

^g^ Department of Respiratory Medicine, North Tees & Hartlepool NHS Foundation Trust, Stockton-on-Tees, United Kingdom

^h^ University Hospitals Birmingham NHS Foundation Trust and Institute of Microbiology & Infection, University of Birmingham, Birmingham, United Kingdom

^i^ University of Manchester and Manchester University NHS Foundation Trust, Manchester, United Kingdom

^j^ James Cook University Hospital, Middlesbrough, United Kingdom

^k^ North West Anglia NHS Foundation Trust, Peterborough, United Kingdom

^l^ Department of Infectious Diseases, Cardiff and Vale University Health Board; Division of Infection and Immunity, Cardiff University, Cardiff, United Kingdom

^m^ Roslin Institute, University of Edinburgh, Edinburgh, United Kingdom

^n^ School of Life Course Sciences, King’s College London, London, United Kingdom

° NIHR Southampton Clinical Research Facility and Biomedical Research Centre, University Hospital Southampton NHS Foundation Trust and University of Southampton, Southampton, United Kingdom

^p^ Department of Mathematics and Statistics, Lancaster University, Lancaster, United Kingdom

^q^ MRC Biostatistics Unit, University of Cambridge, Cambridge, United Kingdom

^r^ Oxford University Hospitals NHS Foundation Trust, Oxford, United Kingdom

^s^ Respiratory Medicine Department, Nottingham University Hospitals NHS Trust, Nottingham, United Kingdom

^t^ School of Medicine, University of Nottingham, Nottingham, United Kingdom

^u^ Intensive Care National Audit & Research Centre, London, United Kingdom

^v^ Mahidol Oxford Tropical Medicine Research Unit, Faculty of Tropical Medicine, Mahidol University, Bangkok, Thailand

^w^ Centre for Tropical Medicine and Global Health, Nuffield Department of Medicine, University of Oxford, United Kingdom

^x^ NIHR Oxford Biomedical Research Centre, Oxford University Hospitals NHS Foundation Trust, Oxford, United Kingdom

*,^†^ equal contribution

### Data Monitoring Committee

Peter Sandercock, Janet Darbyshire, David DeMets, Robert Fowler, David Lalloo, Ian Roberts, Janet Wittes.

## Data Availability

The protocol, consent form, statistical analysis plan, definition & derivation of clinical characteristics & outcomes, training materials, regulatory documents, and other relevant study materials are available online at www.recoverytrial.net
This is a preliminary report and follow-up is ongoing. Data will be made available to bona fide researchers registered with an appropriate institution within 3 months of the final participant completing 28-day follow-up. As described in the protocol, the trial Steering Committee will facilitate the use of the study data and approval will not be unreasonably withheld. However, the Steering Committee will need to be satisfied that any proposed publication is of high quality, honours the commitments made to the study participants in the consent documentation and ethical approvals, and is compliant with relevant legal and regulatory requirements (e.g. relating to data protection and privacy). The Steering Committee will have the right to review and comment on any draft manuscripts prior to publication.

https://www.ndph.ox.ac.uk/data-access

https://www.recoverytrial.net

## Acknowledgements

We would like to thank the many thousands of doctors, nurses, pharmacists, other allied health professionals, and research administrators at 176 NHS hospital organizations across the whole of the UK, supported by staff at the NIHR Clinical Research Network, NHS DigiTrials, Public Health England, Department of Health & Social Care, the Intensive Care National Audit & Research Centre, Public Health Scotland, National Records Service of Scotland, the Secure Anonymised Information Linkage (SAIL) at University of Swansea, and the NHS in England, Scotland, Wales and Northern Ireland. We would especially like to thank the members of the independent Data Monitoring Committee. But above all, we would like to thank the thousands of patients who participated in this study.

## Funding

The RECOVERY trial is supported by a grant to the University of Oxford from UK Research and Innovation/National Institute for Health Research (NIHR) (Grant reference: MC_PC_19056) and by core funding provided by NIHR Oxford Biomedical Research Centre, Wellcome, the Bill and Melinda Gates Foundation, the Department for International Development, Health Data Research UK, the Medical Research Council Population Health Research Unit, the NIHR Health Protection Unit in Emerging and Zoonotic Infections, and NIHR Clinical Trials Unit Support Funding. TF is supported by the NIHR Manchester Biomedical Research Centre. TJ received funding from UK Medical Research Council (MC_UU_0002/14). TJ is supported by a NIHR Senior Research Fellowship (NIHR-SRF-2015-08-001). WSL is supported by core funding provided by NIHR Nottingham Biomedical Research Centre. NJW, JAW and JT are part of the Mahidol Oxford Research Unit supported by the Wellcome Trust. Tocilizumab was provided free of charge for this study by Roche Products Limited. AbbVie contributed some supplies of lopinavir-ritonavir for use in the study. Other medication used in the study was supplied from routine National Health Service stock.

The views expressed in this publication are those of the authors and not necessarily those of the NHS, the National Institute for Health Research or the Department of Health and Social Care (DHCS).

## Conflicts of interest

The authors have no conflict of interest or financial relationships relevant to the submitted work to disclose. No form of payment was given to anyone to produce the manuscript. All authors have completed and submitted the ICMJE Form for Disclosure of Potential Conflicts of Interest. The Nuffield Department of Population Health at the University of Oxford has a staff policy of not accepting honoraria or consultancy fees directly or indirectly from industry (see https://www.ndph.ox.ac.uk/files/about/ndph-independence-of-research-policy-jun-20.pdf).

## Notes

### Competing Interest Statement

The authors have declared no competing interest.

### Clinical Trial

NCT04381936

### Clinical Protocols

https://www.recoverytrial.net

### Author Declarations

The trial was conducted in accordance with the principles of the International Conference on Harmonization Good Clinical Practice guidelines and approved by the UK Medicines and Healthcare Products Regulatory Agency (MHRA) and the Cambridge East Research Ethics Committee (ref: 20/EE/0101).

